# CoviVac vaccination induces production of neutralizing antibodies against Delta and Omicron variants of SARS-CoV-2

**DOI:** 10.1101/2022.02.10.22270781

**Authors:** Liubov Kozlovskaya, Ilya Gordeychuk, Anastasia Piniaeva, Anastasia Kovpak, Anna Shishova, Aleksandr Lunin, Elena Shustova, Vasiliy Apolokhov, Ksenia Fominykh, Yuri Ivin, Alla Kondrashova, Viktor Volok, Irina Tcelykh, Aleksandra Siniugina, Aydar Ishmukhametov

## Abstract

Vaccines are proven to be an effective tool in prophylaxis of severe COVID-19, but emerging mutated SARS-CoV-2 variants constantly challenge vaccines’ protectivity. We have evaluated the ability of the sera from individuals vaccinated with two variants of inactivated vaccine CoviVac and COVID-19 convalescents (May-December 2020) to neutralize SARS-CoV-2 variants Delta and Omicron. Four groups of serum samples (CoviVac vaccinees; COVID-19 convalescents; mice immunized with CoviVac preparations based on prototype B.1.1 strain and Delta variant) were evaluated in virus neutralization test against SARS-CoV-2 heterologous B.1.1 virus, Delta and Omicron variants. CoviVac preparations based on B.1.1 and Delta induced neutralizing antibodies against SARS-CoV-2 B.1.1 and two variants of concern. We observed a decrease in neutralization capacity in the sera from CoviVac (based on B.1.1 strain) vaccinees: 57.1% samples had detectable neutralizing antibodies against Delta and 61.9% against Omicron variants. Sera samples of all (100%) mice immunized with a candidate vaccine based on the SARS-CoV-2 Delta variant strain had neutralizing antibodies against all tested strains.

## 1. Introduction

The severe acute respiratory syndrome coronavirus-2 (SARS-CoV-2) was identified in Wuhan, China, at the end of 2019. The virus belongs to the *Betacoronavirus* genus of *Coronaviridae* family [1]. Since 2019, SARS-CoV-2 has evolved substantially. A number of SARS-CoV-2 variants of interest (VOI) and variants of concern (VOC) have emerged, such as B.1.1.7 (Alpha) lineage that arose in the United Kingdom, B.1.351 (Beta) in South Africa, P.1 (Gamma) in Brazil, B.1.617.2 (Delta) in India and B.1.1.529 (Omicron) all over the world [2]. These variants carry multiple mutations in the receptor-binding domain (RBD) and N-terminal domain (NTD) regions of the main surface virion S (spike) protein [3]. The currently spreading VOC Omicron contains 32 mutations in S protein [4], which drastically change its properties, like transmissibility, pathogenicity, and immune evasion.

Multiple vaccines based on different platforms have been developed to prevent severe COVID-19, generally showing good safety and varying degrees of efficacy [5]. The emergence of variants that may escape from the immune response has raised concerns [6] regarding the decrease of effectiveness of available vaccines and the threat of increased number of re-infections.

Here, we have evaluated the ability of the sera from COVID-19 convalescents (Russia, May-December 2020) and individuals vaccinated with two variants of inactivated vaccine CoviVac to neutralize SARS-CoV-2 variants Delta and Omicron in comparison with prototype strain of B.1.1. lineage.

## 2. Materials and Methods

### 2.1. Cells and viruses

Vero cell line was obtained from Biologicals, World Health Organization, Switzerland. Cells were maintained in Dulbecco’s Modified Eagle Medium (DMEM, Chumakov FSC R&D IBP RAS, Russia), supplemented with fetal bovine serum (FBS, Gibco, 5%), streptomycin (0.1 mg/ml), and penicillin (100 units/ml) (PanEco, Russia).

The SARS-CoV-2 prototype strain PIK35 (Pango lineage B.1.1, GISAID EPI_ISL_428852), variant Delta strain 4724d (GISAID EPI_ISL_8799478) and variant Omicron strain 7995o (GISAID EPI_ISL_9613539) were isolated from nasopharyngeal swab samples of COVID-19 patients in Moscow, Russia. After 3-5 passages in Vero cells, viruses were stored as infected cell suspension at –70°C.

### 2.2. Serum samples

Serum samples were collected from Phase I/II Clinical trials participants, who tested negative for SARS-CoV-2 antibodies in NT and in ELISA before the trials (ClinicalTrials.gov, NCT05046548) and were vaccinated with CoviVac (0.5 ml) intramuscularly twice with 14 days period. Serum samples were collected 7-21 days after the last immunization for vaccine immunogenicity testing. The samples were then stored at –20°C and used in the present study.

COVID-19 convalescent sera samples were collected in Russia from May to December 2020 from convalescent donors according to the state program of plasma collection to be used for treatment.

BALB/c mice were vaccinated with CoviVac preparation based on B.1.1 strain (GISAID EPI_ISL_428851) intramuscularly (0.5 ml) twice with 14 days interval, serum samples were collected 7 days after the last immunization for vaccine immunogenicity testing as described previously [7]. The samples were then stored at –20°C and used in the present study.

The CoviVac candidate vaccine based on Delta variant was prepared using the Delta variant strain (GISAID EPI_ISL_8799478) by the same technology as CoviVac [7]. BALB/c mice were vaccinated with the candidate vaccine (0.5 ml) intramuscularly twice with 14 days interval, serum samples were collected 7 days after the last immunization for vaccine immunogenicity testing. The samples were then stored at –20°C and used in the present study.

### 2.3. Neutralization test (NT)

The NT was described previously [7]. In brief, eight two-fold serum dilutions were prepared in DMEM (Chumakov FSC R&D IBP RAS, Russia). The dilutions were mixed with equal volumes of virus suspension containing 50-200 TCID50 of the respective viruses per well. Final serum dilution series started from 1:8. After 1 h incubation at 37°C, virus-serum mixtures were added to the confluent Vero cell monolayers in 2 replicates. In parallel with every test run, control cells were incubated with the analogous sequential dilutions of non-immune and standard immune control sera; and a virus dose titration was performed. After a 5-day incubation at 37 °C, cytopathic effect (CPE) was visually assessed via a light microscope. Neutralizing antibody (nAB) titers were calculated according to the Karber method [8]. The result of <1:8 was considered negative.

### 2.4. Statistical analysis

Data series were tested for normality using Shapiro-Wilk test. Most data series were normally distributed; therefore, Student T-test was used to compare values. Comparisons and visualizations were performed in OriginPro 8 (OriginLab, USA).

## 3. Results and Discussion

Several studies have shown significant decrease in neutralization of novel VOCs, especially variant Omicron, by the sera from individuals immunized with various vaccines used in various combinations. A standard two-shot course of vaccination with mRNA vaccines mRNA-1273 [9] and BNT162b2 [10] induced detectable antibodies against variant Omicron in 85% and 38.2% of vaccine recipients, respectively. For inactivated vaccine CoronaVac, nABs against Omicron were not detected in 25 samples with aABs to the ancestral lineage A virus [11]. For another inactivated vaccine BBIBP-CorV in the two-dose vaccination group nAB levels against Omicron were below the lower limit of quantitation in 80% of the samples 14 days following the second dose. Nevertheless, after homological or heterological booster vaccination 100% samples showed positive neutralization activity against Omicron [12].

Serum samples from four groups were assessed in this study: CoviVac clinical trials participants; mice immunized with CoviVac for vaccine immunogenicity testing (as a part of the production process); mice immunized with candidate CoviVac preparation based on Delta variant strain; and COVID-19 convalescents (Moscow, Russia, May-December 2020). Serum samples were previously tested in NT against SARS-CoV-2 B.1.1 lineage strain PIK35. Thus, only positive samples were selected with nAB titers of 4.5log2 (1:23) – 8log2 (1:256), approximately 20 samples per group. Samples collected from COVID-19 convalescent donors had higher nAB titers against the B.1.1 virus, 6.5log2 (1:91) – 8log2 (1:256).

Serum samples from all groups were simultaneously tested (in the same neutralization test experiment) against SARS-CoV-2 Delta variant strain 4724d and Omicron variant strain 7995o. The results are presented in Table 1 and Figure 1.

**Table 1.** Proportion of serum samples with detectable nABs against SARS-CoV-2 B.1.1 lineage strain PIK35, Delta variant strain 4724d and Omicron variant strain 7995o in selected serum samples of four groups.

| Serum samples | No. positive in NT/total No. (%) |  |  |
| --- | --- | --- | --- |
|  | B.1.1<br>(st. PIK35) | Delta<br>(st. 4724d) | Omicron<br>(st. 7995o) |
| CoviVac vaccinated humans | 21/21 (100%) | 12/21 (57.1%) | 13/21 (61.9%) |
| CoviVac vaccinated mice | 16/16 (100%) | 10/16 (62.5%) | 16/16 (100%) |
| Candidate CoviVac (from Delta variant strain) vaccinated mice | 20/20 (100%) | 20/20 (100%) | 20/20 (100%) |
| COVID-19 convalescents | 20/20 (100%) | 20/20 (100%) | 20/20 (100%) |

Serum samples with nAB titer <1:8 were considered negative

**Figure 1.**
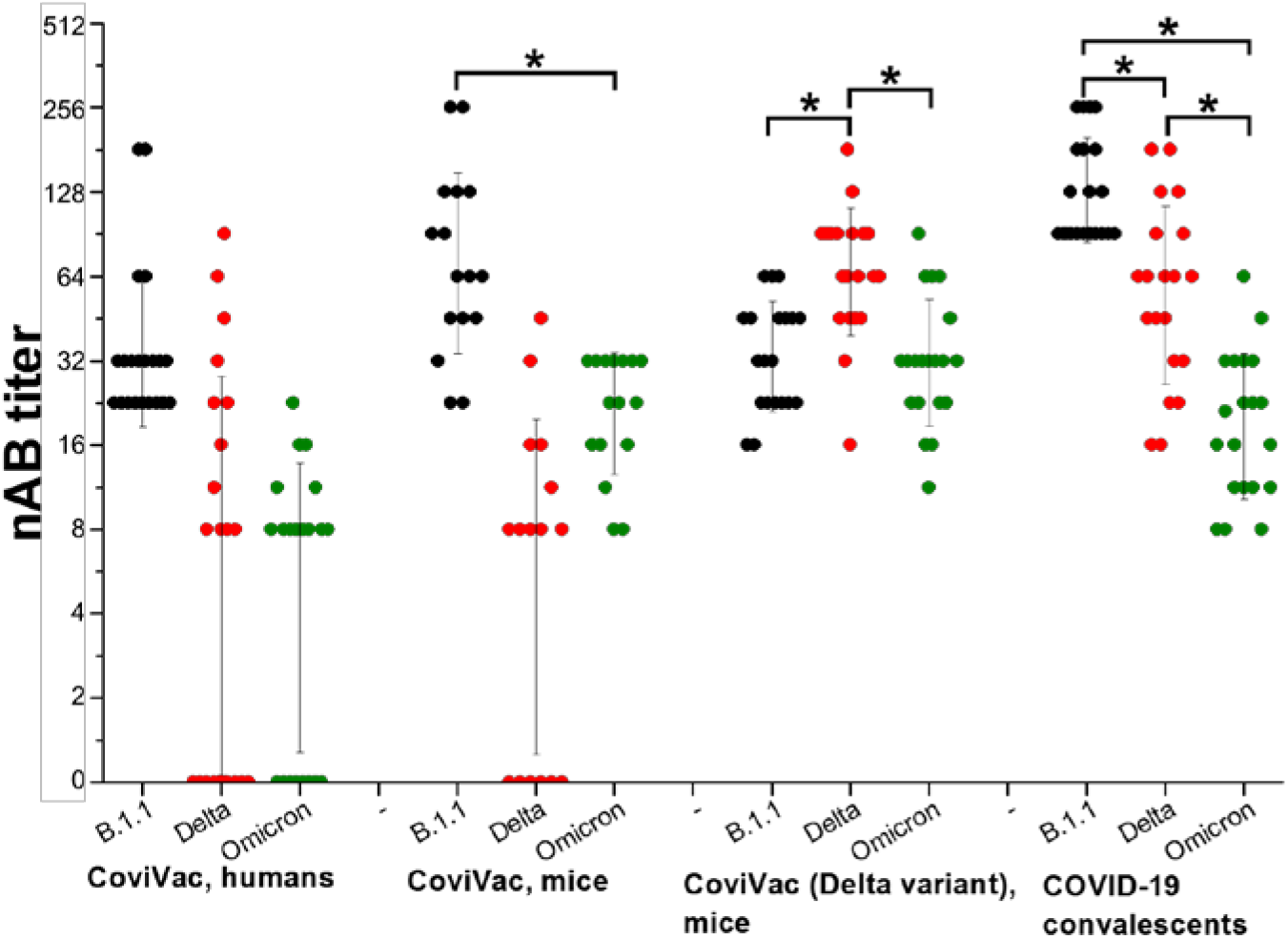
Serum neutralization tests (NTs) against SARS-CoV-2 B.1.1 lineage strain PIK35, Delta variant strain 4724d and Omicron variant strain 7995o. Serum samples with nAB titer <1:8 were approximated as 0log2. * – difference is statistically significant (T test, p < 0.05)

Serum samples with comparatively high nAB titers collected from COVID-19 convalescents successfully neutralized both VOCs. However, the nAB titers against VOCs were significantly lower, which supports the previously published data.

Serum samples collected from CoviVac clinical trials participants showed reduced neutralization with Delta and Omicron variants of SARS-CoV-2. nABs were present in detectable titers (>1:8) in 12/21 (57.1%) and 13/21 (61.9%) of the serum samples, respectively. The results were supported by the data obtained in immunized mice serum samples, where nABs were found in detectable titers (>1:8) in 10/16 (57.1%) and 16/16 (100%) samples, respectively. The difference is most likely due to the overall higher titers in the tested mouse samples. Overall, these data correlate with the other neutralization studies [9,10]. A relatively higher proportion of serum samples with nABs against variant Omicron in CoviVac recipients compared to some other inactivated vaccines may be due to the presence of D614G mutation in the vaccine strain.

Serum samples of all (100%) mice vaccinated with the candidate vaccine CoviVac based on the SARS-CoV-2 Delta variant strain had detectable nABs against B.1.1 strain PIK35 and Omicron variant strain 7995o. Still, a decrease in nAB titers was observed in both cases. The presence of nABs to all three SARS-CoV-2 variant strains in sera of mice immunized with the candidate vaccine based on Delta strain can be explained by the fact that Delta variant S protein contains several mutations common with the Omicron variant. Moreover, the neutralization levels against a B.1.1 strain for candidate CoviVac from Delta variant are still very high.

## 4. Conclusions

Inactivated vaccine CoviVac and candidate vaccine CoviVac (based on Delta variant strain) induced neutralizing antibodies against SARS-CoV-2 prototype B.1.1 strain and two VOCs. However, in the sera from CoviVac vaccinees a decrease in nAB titers against strains of Delta and Omicron VOCs were observed. The switch of the vaccine strain to Delta variant one may improve the efficacy of the inactivated vaccine. The effect of booster immunization is to be evaluated in further studies.

## Data Availability

All data produced in the present work are contained in the manuscript

## Author Contributions

Conceptualization, L.K., I.G., Al.S. and A.I.; methodology, L.K., I.G., and An.S.; investigation, L.K., A.K., An.S., A.L., E.S., V.A., K.F., Y.I., V.V., I.T.; writing—original draft preparation, L.K..; writing—review and editing, A.K. and I.G.; visualization, L.K.; supervision, A.P. and Al.S.; project administration, A.K.; funding acquisition, A.I. All authors have read and agreed to the published version of the manuscript.

## Informed Consent Statement

All participants provided written informed consent before inclusion in the Clinical trials study, which cover the use of the serum samples for vaccine immunogenicity testing.

## Data Availability Statement

The data generated in this study is present on the main text in full.

## References

1. ICTV [Electronic resource]. URL: https://talk.ictvonline.org/taxonomy/ (accessed: 04.02.2022).

2. World Health Organization. Tracking SARS-CoV-2 variants // Who. 2021. P. https://www.who.int/en/activities/tracking-SARS-Co.

3. SARS-CoV-2 variants ∼ ViralZone [Electronic resource]. 2021. URL: https://viralzone.expasy.org/9556 (accessed: 04.02.2022).

4. He X. et al. SARS-CoV-2 Omicron variant: Characteristics and prevention // MedComm. MedComm (2020), 2021. Vol. 2, №4. P. 838–845.

5. Yan Z., Yang M., Lai C.L. COVID-19 Vaccines: A Review of the Safety and Efficacy of Current Clinical Trials // Pharmaceuticals (Basel). Pharmaceuticals (Basel), 2021. Vol. 14, №5.

6. Li M., Lou F., Fan H. SARS-CoV-2 variant Omicron: currently the most complete “escapee” from neutralization by antibodies and vaccines // Signal Transduct. Target. Ther. Nature Publishing Group, 2022. Vol. 7, №1. P. 28.

7. Kozlovskaya L.I. et al. Long-term humoral immunogenicity, safety and protective efficacy of inactivated vaccine against COVID-19 (CoviVac) in preclinical studies // Emerg. Microbes Infect. Emerg Microbes Infect, 2021. Vol. 10, №1. P. 1790–1806.

8. Kärber G. Beitrag zur kollektiven Behandlung pharmakologischer Reihenversuche // Naunyn. Schmiedebergs. Arch. Exp. Pathol. Pharmakol. Springer, 1931. Vol. 162, №4. P. 480–483.

9. Pajon R. et al. SARS-CoV-2 Omicron Variant Neutralization after mRNA-1273 Booster Vaccination // N. Engl. J. Med. N Engl J Med, 2022.

10. Chen L.-L. et al. Omicron variant susceptibility to neutralizing antibodies induced in children by natural SARS-CoV-2 infection or COVID-19 vaccine // Emerg. Microbes Infect. Emerg Microbes Infect, 2022. P. 1–17.

11. Lu L. et al. Neutralization of Severe Acute Respiratory Syndrome Coronavirus 2 Omicron Variant by Sera From BNT162b2 or CoronaVac Vaccine Recipients // Clin. Infect. Dis. Oxford University Press, 2021.

12. Ai J. et al. Omicron variant showed lower neutralizing sensitivity than other SARS-CoV-2 variants to immune sera elicited by vaccines after boost // Emerg. Microbes Infect. Emerg Microbes Infect, 2022. Vol. 11, №1. P. 337–343.

